# Clinical and pathological characteristics of thin cutaneous melanomas with rapid recurrence

**DOI:** 10.64898/2026.04.04.26350182

**Authors:** Prachi Bhave, Terence Wong, Kim Margolin, Lotte Hoeijmakers, Johanna Mangana, Maria Grazia Vitale, Paolo A Ascierto, Andrea Maurichi, Mario Santinami, Georgina Heddle, Clara Allayous, Celeste Lebbe, Adnan Kattak, Stephan Forchhammer, Jolien I Kessels, Peter Lau, Serigne N Lo, Anthony A Papenfuss, Grant A McArthur

## Abstract

**Background:** Although thin, T1 melanomas have an excellent cure rate with surgery alone, >25% of melanoma deaths originate from thin melanomas (TMs). There is, therefore, an urgent need to improve the identification and management of patients with TMs at high risk of recurrence.

**Methods:** Patients with T1 melanoma and recurrence ≤ 2 years of diagnosis (T1 rapid group) were compared to patients with T1 melanoma and recurrence ≥10 years after diagnosis (T1 late group).

**Results:** 442 patients from 14 sites were included: 310 and 132 patients in the T1 rapid and late groups, respectively. Median age at primary melanoma diagnosis was 51 years [15-85], 272 (62%) male, 254 (58%) superficial spreading and 101 (23%) head/neck primary. The majority (73%) of recurrences in the T1 rapid group were locoregional. Using univariable logistic regression analysis, age >65 years (p<0.0001), lentigo maligna (LM) melanoma subtype (p=0.025), head/neck primary site (p=0.0065), mitoses ≥1/mm^2^ (p=0.0181) and ulceration (p=0.0087) were significantly associated with T1 rapid recurrence compared to T1 late recurrence. Using multivariable analysis, age >65 years (p=0.0010), mitoses ≥1/mm^2^ (p=0.049) and ulceration (p=0.037) remained significant.

**Conclusions:** Rapid recurrence of TM is associated with age >65 years, LM subtype, head/neck primary site, mitoses ≥1/mm^2^ and ulceration.

## Background

Melanoma incidence has been rising over the last 50 years in fair-skinned populations of European ancestry, with most of this increase being due to more thin melanomas being diagnosed.^1^ The incidence of early-stage melanoma is so high that more patients are diagnosed with thin, stage I melanoma than all other stages combined.^1,2^ Melanoma survival rates correlate strongly with thickness of the primary skin lesion, with patients with thicker melanomas having substantially higher risk of mortality.^2^ Thin, stage I melanomas have an excellent chance of cure with surgery alone, with only ∼2% recurring and leading to patient death within 10 years of diagnosis of the primary.^3^ Longer follow-up has also demonstrated that 30-year melanoma-specific survival rates of patient with stage I melanoma are almost 95%.^4,5^ Despite this overall favourable prognosis, thin, early-stage melanomas account for over 25% of melanoma deaths, because of the sheer volume of such cases.^2,3,6^ Moreover, studies have shown that more patients die from thin melanomas than thick melanomas.^6,7^ There is, therefore, an urgent need to improve the identification of patients with early-stage melanoma at high risk of recurrence.

Despite the increasing incidence of melanoma, mortality rates from advanced melanoma have decreased significantly over the last decade in some countries such as Australia, largely due to the advent of novel systemic therapies such as checkpoint-inhibiting immunotherapies and BRAF/MEK targeted agents.^8^ These treatments have resulted in significant improvements in patient survival rates for those with stage IV (distant metastatic) melanoma, with a 50% reduction in age standardised mortality.^9,10^ These agents have also halved the recurrence rates of stage III (regionally or local metastatic) melanoma when given as adjuvant therapy, after surgical removal of the melanoma.^11,12^ Recent studies have also shown an improvement in patient outcomes when adjuvant immunotherapy is used in patients with high-risk stage II melanoma.^13,14^ Furthermore, neoadjuvant immunotherapy, administered prior to the surgical removal of certain stage III melanoma, has very recently been shown to provide significant improvements in event-free survival.^15^

Identifying the clinical and molecular features of thin, stage I melanomas that confer poor prognosis is increasingly a focus of research studies. The prospect of treating such patients with adjuvant therapy has led to various strategies to help identify patients with thin melanoma who are at high risk of death, such as through gene expression profiling (GEP), sentinel lymph node biopsy (SLNB) and prognostic models.^16–19^ Furthermore, studies have shown that thin melanomas are more likely to recur late, over 5 years from diagnosis.^4,20^ This makes diagnosis of recurrence and patient management even more challenging, as patients have often completed follow up or surveillance programs well before disease recurrence.

This study examines the clinical and pathological features of patients with early-stage, thin melanoma and unexpected clinical outcomes. Patients with thin, T1 melanoma (≤1mm thickness) and rapid recurrence of melanoma within 2 years of diagnosis are compared to patients with T1 melanoma and late recurrence, 10 years after diagnosis. A threshold of 2 years since diagnosis for the rapid recurrence cohort was chosen due to the significant clinical impact of such an early recurrence, and is also based on the hypothesis that such rapidly recurring tumours are likely to have distinct underlying biology compared to those with later recurrence.^21.22^ A threshold of 10 years after diagnosis for the late recurrence group was chosen as it is known that a large number of deaths from T1 melanoma occur after 10 years.^4^

## Methods

### Patients and study design

Following institutional ethics board approval (Peter Mac No 07/38), consecutive patients meeting one of the following two criteria were identified retrospectively: i) T1 melanoma with recurrence within 2 years of diagnosis ii) T1 melanoma with recurrence at least 10 years after diagnosis. Melanoma was staged per the American Joint Committee on Cancer (AJCC) eighth edition staging system.^23^ Data on patient, disease and treatment characteristics were collected, including: patient demographics (age, sex, treating centre, comorbidities, history of chronic sun damage, ethnicity, skin type, family history of melanoma); primary melanoma characteristics (Breslow thickness, ulceration, mitosis, regression, tumour infiltrating lymphocytes (TILs), lymphovascular invasion (LVI), associated naevus, histopathological subtype, primary location, genomic mutation(s), method of diagnosis); first recurrence details (method of detection, site and stage of recurrence, treatment), number of further recurrences and survival outcomes. The Fitzpatrick classification was used to categorise patient skin type. Radiological response to systemic therapy for first recurrence was assessed by Response Evaluation Criteria in Solid Tumors (RECIST V.1.1).^24^ Best response to adjuvant therapy after surgery for first recurrence was considered progressive disease (PD) if recurrence occurred during or within 6 months of completion of adjuvant therapy. Oncogene mutation testing may have been performed on the primary melanoma or recurrent disease. Patients with multiple primary melanomas were excluded. Patients were followed until death or data censorship date, whichever occurred first.

Time to first recurrence was calculated from date of primary melanoma diagnosis as confirmed by initial biopsy to date of radiological or clinical first recurrence. Recurrence free survival (RFS) was calculated from date of primary melanoma diagnosis to date of radiological or clinical first recurrence, death or last follow-up. Overall survival (OS) was calculated from date of primary melanoma diagnosis to date of death from any cause or last follow-up.

### Statistical analysis

Data were described overall and stratified by group. Categorical variables were summarised as frequencies and percentages, and groups difference compared using Pearson’s χ2 test and/or Fisher’s exact test, as appropriate. Continuous variables were described using median (range) and differences between group were assessed using the Kruskal-Wallis test. Univariable and multivariable logistic regression analyses were conducted to investigate factors associated with first recurrence in the two patient subgroups. Variables that were statistically significant in the univariable analysis or considered to be clinically relevant were included in the multivariable analysis. RFS was analysed using univariable and multivariable Cox proportional hazard regression. RFS and OS were described using the Kaplan-Meier method and examined for both patient subgroups, reporting median survival time with 95% confidence intervals (CI). Differences between survival curves were assessed using the log-rank test. Characteristics with more than 25% missing data were excluded from the chi-square test and subsequent analyses. All tests were two-sided, with a p-value of <0.05 considered statistically significant. All statistical analyses were conducted using SAS 9.4 (SAS Institute Inc. Cary, NC, USA) and R 4.3.1 (R Core Team, Vienna, Austria).

## Results

A total of 442 patients from fourteen sites in Australia, Europe and the USA were included in the final analysis. Of these, 310 patients had T1 melanoma with recurrence within 2 years of diagnosis (‘rapid group’) and 132 patients had T1 melanoma with recurrence more than 10 years after diagnosis (‘late group’). Baseline patient and disease characteristics are summarised in **Table 1**. Characteristics with more than 25% of patients having unknown data are detailed in **Supplement 1**. Across the cohort, median age at diagnosis of primary melanoma was 51 years (range, 15-85), with median age being higher in the T1 rapid group compared to T1 late group (55 years vs 43 years, p<0.001); 272 (62%) were male, 174 (39%) had Fitzpatrick skin type II, 356 (81%) had no family history of melanoma and 129 (29%) had at least 1 comorbidity. For both T1 groups, median melanoma Breslow thickness (BT) was 0.8mm and mean BT was 0.7mm. Several patients (59, 13%) had very thin melanoma of less than 0.5mm BT. Most melanomas were non-ulcerated (302, 68%) and had a mitotic rate of 0 mitoses/mm^2^ (362,82%). The majority (58%) of patients had superficial spreading melanoma (SSM) and most (46%) melanomas originated from the trunk, back or abdomen. Of the entire cohort, 103 (23%) patients had oncogene mutation testing performed; of these patients, 45 (44%) had a BRAF V600 mutation, consistent with the expected prevalence of BRAF V600 mutations in an unselected population of patients with cutaneous melanoma. Approximately 13% of the cohort underwent sentinel lymph node biopsy (SLNB), equally split between the T1 rapid and T1 late groups, though patients with T1b melanoma were more likely to undergo SLNB than patients with T1a melanoma (**Table 1**).

**Table 1:**
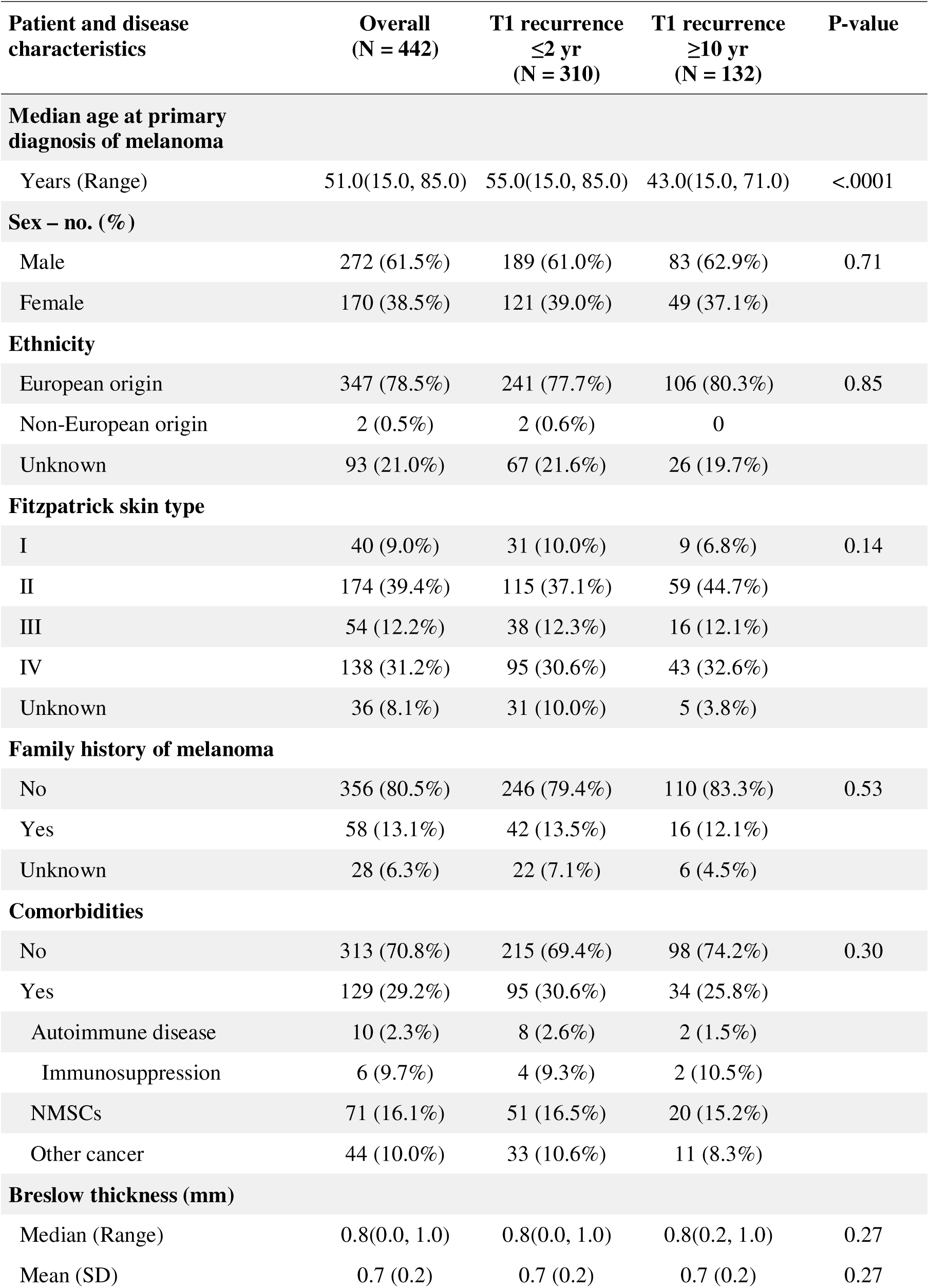
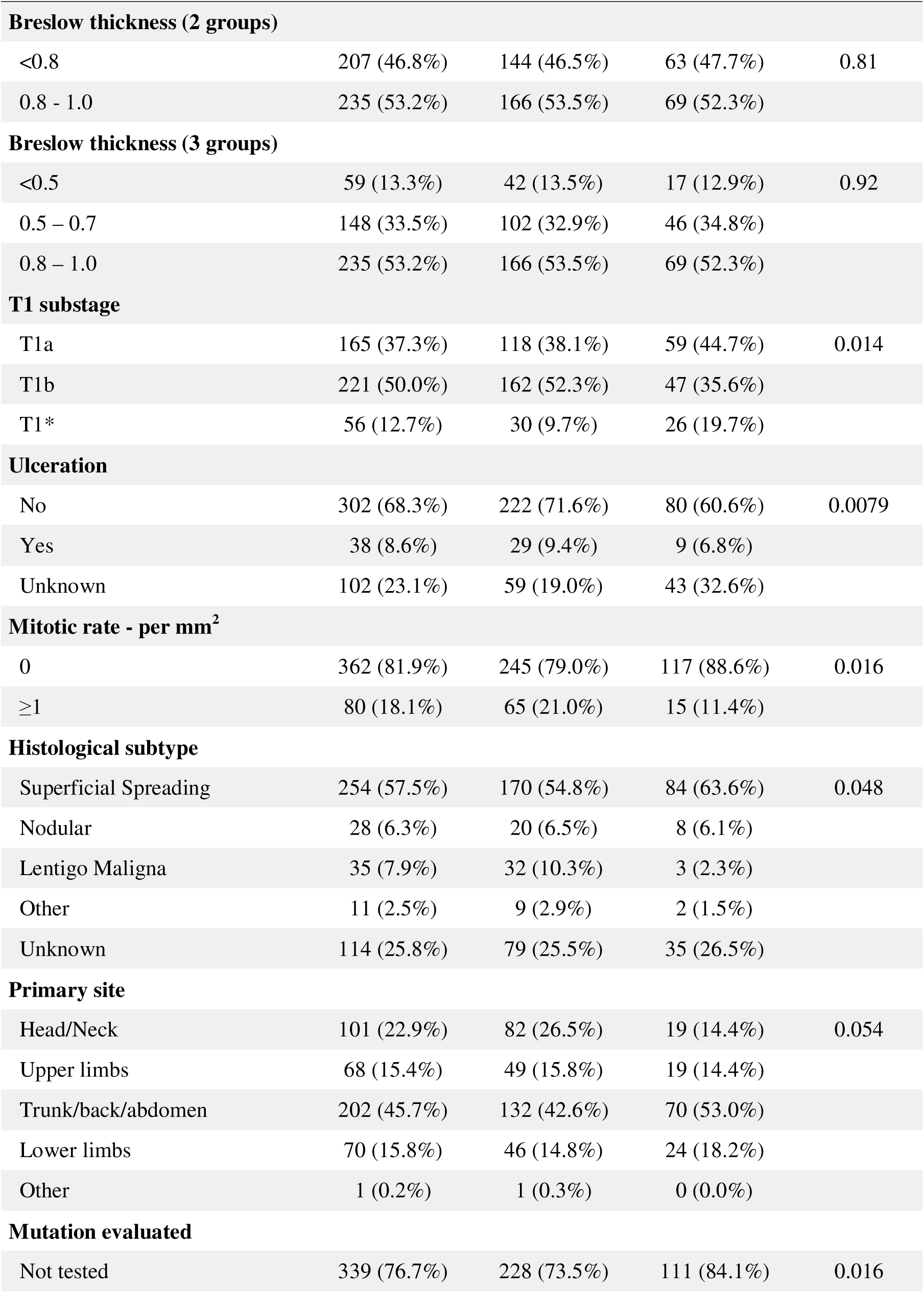
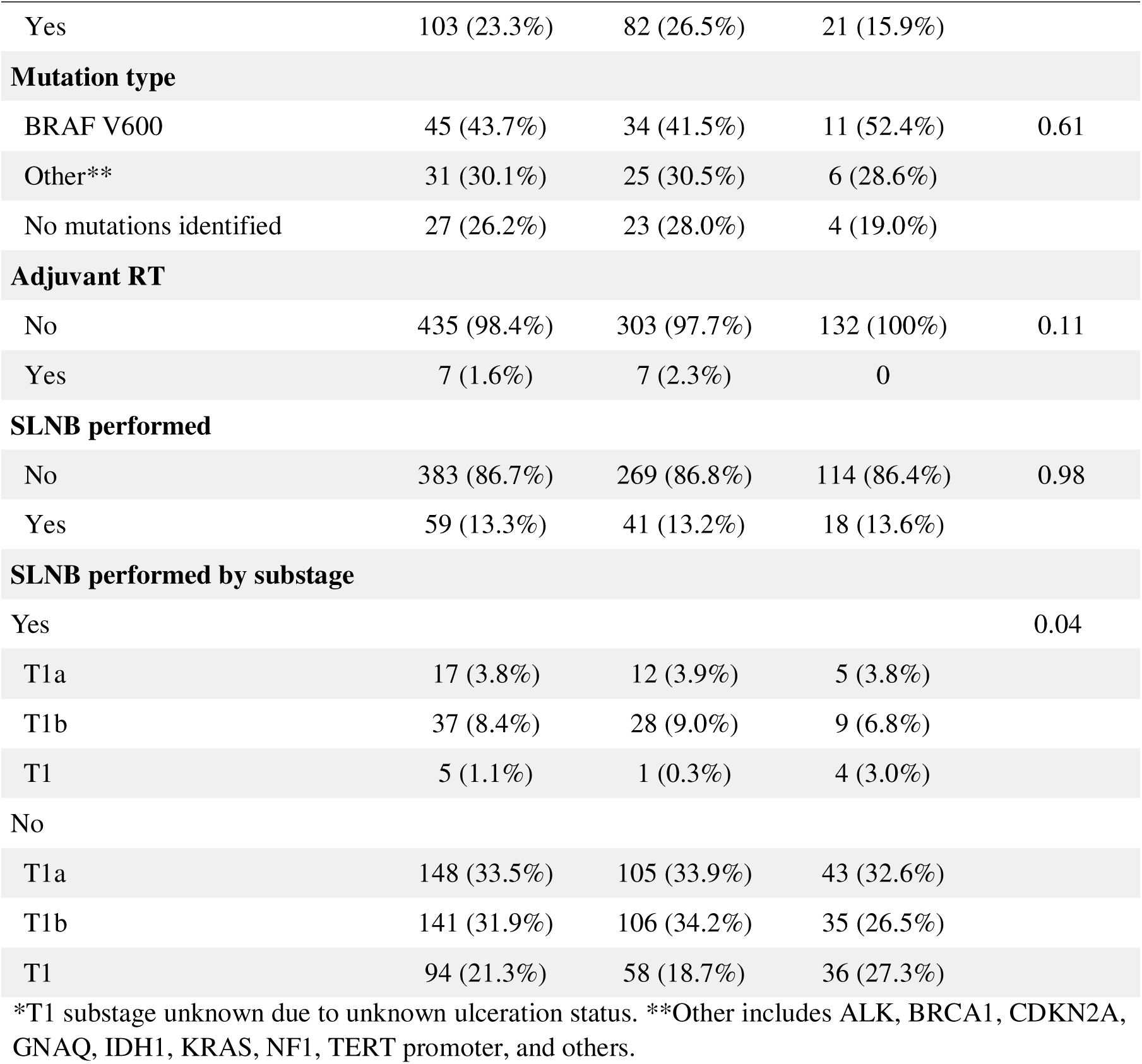
Baseline characteristics for all patients and stratified by recurrence groups. NMSCs, non-melanoma skin cancers; RT, radiotherapy; SLNB, sentinel lymph node biopsy.

Patient and disease characteristics at first recurrence are summarised in **Table 2**. Median time to first recurrence was 1.2 years for the T1 rapid group and 13.6 years for the T1 late group. Where method of detection of recurrence was recorded, most (74/146, 51%) patients in the T1 rapid group presented with symptoms whilst most (23/58, 40%) patients in the T1 late group had their recurrence detected by clinical examination, though there was a large portion (>50%) with unknown method of recurrence detection in both groups. The majority (225/310, 73%) of patients in the T1 rapid group developed locoregional disease at first recurrence, most frequently stage IIIB disease, and recurrence was managed by surgical resection (83/310, 27%) or surgery and adjuvant systemic therapy (+/-radiotherapy) in most (155/310, 50%) patients. Comparatively, first recurrence in the T1 late group most commonly presented with stage IV disease (75/132, 57%), the majority of which was M1c disease and where one site was involved with distant metastases. A variety of systemic therapies were used to treat first recurrence, as outlined in **Table 2**, including both historical treatments such as BCG vaccine and interferon, as well as more contemporary therapies such as checkpoint inhibitors. Most patients across both T1 groups developed further (>1) melanoma recurrences after treatment of the first recurrence.

**Table 2:** Characteristics of first recurrence for all patients and stratified by recurrence groups. All staging per AJCC 8th edition. RT, radiotherapy; BSC, best supportive care; NED, no evidence of disease. *Other treatments included interleukins, tamoxifen, cimetidine, vaccines, transfer factor, NSAIDs and combinations of these.

| Patient and disease characteristics | Overall<br>(N = 442) | T1 recurrence<br>≤2 yr<br>(N = 310) | T1 recurrence<br>≥10 yr<br>(N = 132) | P-value |
| --- | --- | --- | --- | --- |
| **Median time to first recurrence** |  |  |  |  |
| Years (Range) | 1.5(0.1, 32.9) | 1.2(0.1, 2.1) | 13.6(10.1, 32.9) | <.0001 |
| **Method of detection** |  |  |  |  |
| Symptoms | 89 (20.1%) | 74 (23.9%) | 15 (11.4%) | 0.0024 |
| Clinical examination | 74 (16.7%) | 51 (16.5%) | 23 (17.4%) |  |
| Imaging | 41 (9.3%) | 21 (6.8%) | 20 (15.2%) |  |
| Unknown | 238 (53.8%) | 164 (52.9%) | 74 (56.1%) |  |
| **Site of first recurrence** |  |  |  |  |
| Locoregional | 283 (64.0%) | 225 (72.6%) | 58 (43.9%) | <.0001 |
| Distant | 127 (28.7%) | 63 (20.3%) | 64 (48.5%) |  |
| Both | 32 (7.2%) | 22 (7.1%) | 10 (7.6%) |  |
| **Stage at first recurrence** |  |  |  |  |
| III | 279 (63.1%) | 222 (71.6%) | 57 (43.2%) | <.0001 |
| IV | 163 (36.9%) | 88 (28.4%) | 75 (56.8%) |  |
| **Stage III substage** |  |  |  |  |
| IIIA | 22 (7.9%) | 18 (8.1%) | 4 (7.0%) | 0.50 |
| IIIB | 206 (73.8%) | 166 (74.8%) | 40 (70.2%) |  |
| IIIC | 49 (17.6%) | 37 (16.7%) | 12 (21.1%) |  |
| IIID | 2 (0.7%) | 1 (0.5%) | 1 (1.8%) |  |
| **Stage IV, M substage** |  |  |  |  |
| M1a | 29 (17.8%) | 17 (19.3%) | 12 (16.0%) | 0.40 |
| M1b | 38 (23.3%) | 24 (27.3%) | 14 (18.7%) |  |
| M1c | 62 (38.0%) | 32 (36.4%) | 30 (40.0%) |  |
| M1d | 34 (20.9%) | 15 (17.0%) | 19 (25.3%) |  |
| **If stage IV, number of sites involved with distant metastases** |  |  |  |  |
| 1 | 75 (46.0%) | 38 (43.2%) | 37 (49.3%) | 0.88 |
| 2 | 41 (25.2%) | 23 (26.1%) | 18 (24.0%) |  |
| 3 | 25 (15.3%) | 14 (15.9%) | 11 (14.7%) |  |
| >3 | 22 (13.5%) | 13 (14.8%) | 9 (12.0%) |  |
| Management of first recurrence |  |  |  |  |
| Surgery | 119 (26.9%) | 83 (26.8%) | 36 (27.3%) | 0.039 |
| RT | 6 (1.4%) | 1 (0.3%) | 5 (3.8%) |  |
| Systemic Rx | 61 (13.8%) | 44 (14.2%) | 17 (12.9%) |  |
| Surgery + RT | 24 (5.4%) | 18 (5.8%) | 6 (4.5%) |  |
| Surgery + Systemic Rx | 156 (35.3%) | 115 (37.1%) | 41 (31.1%) |  |
| Surgery + RT + systemic Rx | 29 (6.6%) | 22 (7.1%) | 7 (5.3%) |  |
| RT + systemic Rx | 13 (2.9%) | 9 (2.9%) | 4 (3.0%) |  |
| Nil/BSC | 34 (7.7%) | 18 (5.8%) | 16 (12.1%) |  |
| Systemic therapy drug type |  |  |  |  |
| PD1 | 30 (11.6%) | 26 (13.7%) | 4 (5.8%) | 0.26 |
| PD1/CTLA4 | 20 (7.7%) | 12 (6.3%) | 8 (11.6%) |  |
| BRAF/MEKi | 12 (4.6%) | 9 (4.7%) | 3 (4.3%) |  |
| CTLA4 | 7 (2.7%) | 7 (3.7%) | 0 |  |
| Other* | 43 (16.6%) | 29 (15.3%) | 14 (20.3%) |  |
| BCG vaccine | 69 (26.6%) | 51 (26.8%) | 18 (26.1%) |  |
| Interferon | 25 (9.7%) | 19 (10.0%) | 6 (8.7%) |  |
| Chemotherapy | 24 (9.3%) | 19 (10.0%) | 5 (7.2%) |  |
| Unknown | 21 (8.1%) | 12 (6.3%) | 9 (13.0%) |  |
| IO based trial | 8 (3.1%) | 6 (3.2%) | 2 (2.9%) |  |
| Systemic therapy treatment intent |  |  |  |  |
| Adjuvant | 174 (67.2%) | 130 (68.4%) | 44 (63.8%) | 0.62 |
| Palliative | 83 (32.0%) | 58 (30.5%) | 25 (36.2%) |  |
| Unknown | 2 (0.8%) | 2 (1.1%) | 0 |  |
| Best response (Adjuvant) |  |  |  |  |
| Progressive Disease | 60 (46.2%) | 46 (48.4%) | 14 (40.0%) | 0.39 |
| NED | 70 (53.8%) | 49 (51.6%) | 21 (60.0%) |  |
| Best response (Non-adjuvant) |  |  |  |  |
| Progressive Disease | 22 (41.5%) | 16 (40.0%) | 6 (46.2%) | 0.56 |
| Stable Disease | 13 (24.5%) | 11 (27.5%) | 2 (15.4%) |  |

| **Patient and disease characteristics** | **Overall**<br>**(N = 442)** | **T1 recurrence**<br>**≤2 yr**<br>**(N = 310)** | **T1 recurrence**<br>**≥10 yr**<br>**(N = 132)** | **P-**<br>**value** |
| --- | --- | --- | --- | --- |
| Partial Response | 11 (20.8%) | 9 (22.5%) | 2 (15.4%) |  |
| Complete Response | 7 (13.2%) | 4 (10.0%) | 3 (23.1%) |  |
| **Further recurrences** |  |  |  |  |
| No | 146 (40.4%) | 98 (38.7%) | 48 (44.4%) | 0.31 |
| Yes | 215 (59.6%) | 155 (61.3%) | 60 (55.6%) |  |
| 1 | 72 (33.5%) | 54 (34.8%) | 18 (30.0%) | 0.19 |
| 2 | 58 (27.0%) | 42 (27.1%) | 16 (26.7%) |  |
| 3 | 33 (15.3%) | 27 (17.4%) | 6 (10.0%) |  |
| >3 | 52 (24.2%) | 32 (20.6%) | 20 (33.3%) |  |
| **Median follow up time from**<br>**primary melanoma diagnosis** |  |  |  |  |
| Years (Range) | 7.7(0.6, 46.8) | 4.3(0.6, 46.8) | 17.1(10.5, 37.7) | <.0001 |
| **Median follow up time from first**<br>**recurrence** |  |  |  |  |
| Years (Range) | 2.9(0.0, 44.9) | 3.2(0.1, 44.9) | 2.2(0.0, 19.6) | 0.0071 |
| **Survival** |  |  |  |  |
| Death from other cause | 20 (4.5%) | 12 (3.9%) | 8 (6.1%) | 0.23 |
| Death from melanoma | 244 (55.2%) | 164 (52.9%) | 80 (60.6%) |  |
| Cause of death unknown | 18 (4.1%) | 13 (4.2%) | 5 (3.8%) |  |
| Alive | 160 (36.2%) | 121 (39.0%) | 39 (29.5%) |  |

Median follow-up from primary melanoma diagnosis was 4.3 years and 17.1 years for the T1 rapid and T1 late groups, respectively. Death due to melanoma occurred in 164 (53%) and 80 (61%) patients in the T1 rapid and T1 late groups, respectively. Median RFS was 14.1 months (95% CI, 13.0-15.4) and 13.6 years (95% CI, 12.8-14.2) in the T1 rapid and T1 late groups, respectively (**Figure 1**). Median OS was 5.1 years (95% CI, 4.4-6.1) and 18.0 years (95% CI, 17.0-21.8) in the T1 rapid and T1 late groups, respectively (**Figure 2**).

**Figure 1:**
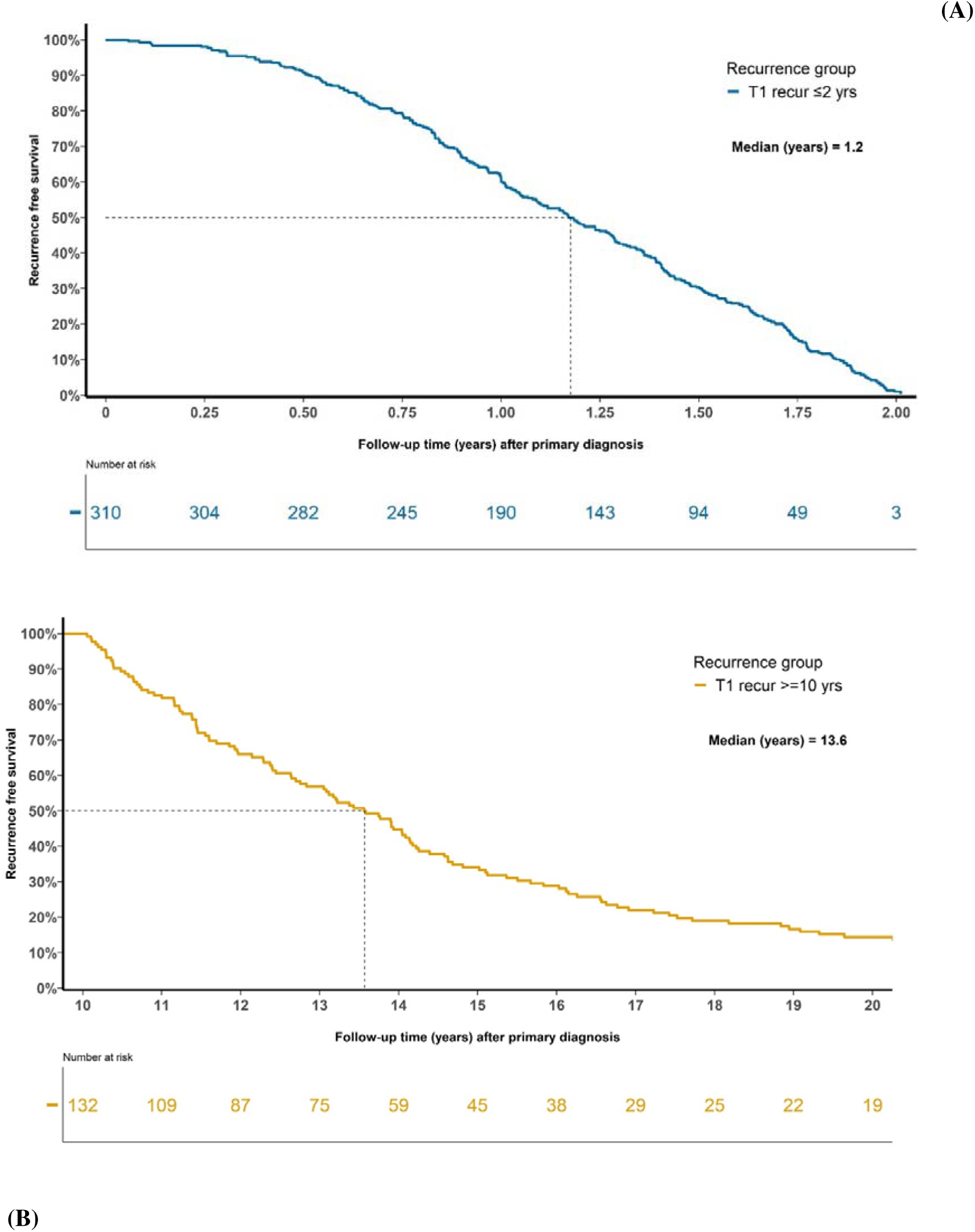
Kaplan-Meier curve for RFS for (A) patients with T1 melanoma and recurrence ≤ 2 years of diagnosis, (B) patients with T1 melanoma and recurrence ≥ 10 years of diagnosis.

**Figure 2:**
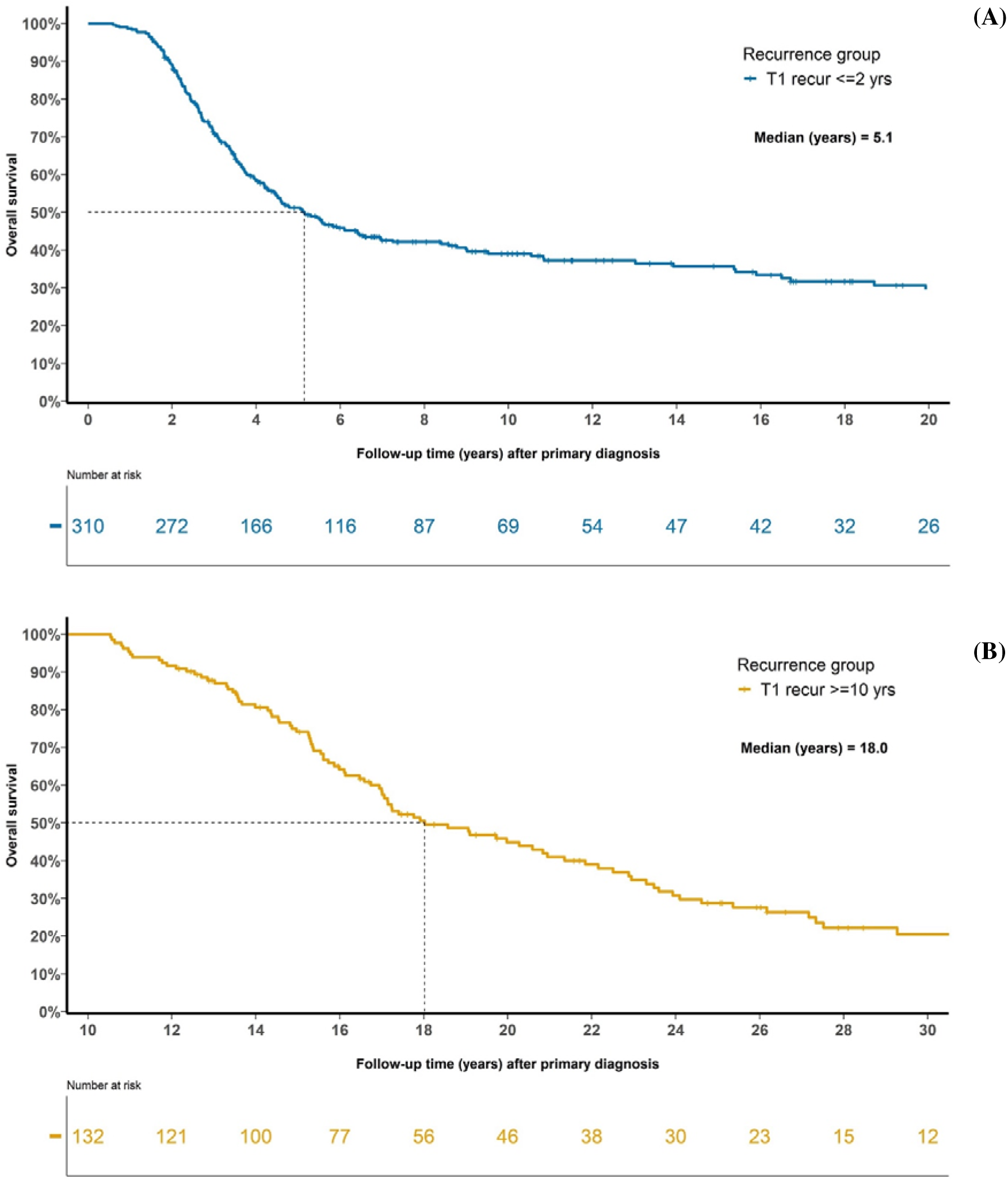
Kaplan-Meier curve for OS for (A) patients with T1 melanoma and recurrence ≤ 2 years of diagnosis, (B) patients with T1 melanoma and recurrence ≥ 10 years of diagnosis.

Univariable logistic regression analysis revealed that age >65 years (p<0.0001), lentigo maligna (LM) melanoma histopathological subtype (p=0.025), head or neck primary melanoma site (p=0.0065), mitoses ≥1/mm^2^ (p=0.0181) and ulceration (p=0.0087) were significantly associated with T1 rapid recurrence compared to T1 late recurrence. On multivariable analysis, age >65 years (HR 3.0, p=0.0009) and ulceration (HR 1.12. p=0.042) remained significantly associated with rapid recurrence, with a trend towards significance for mitoses ≥1/mm^2^ (HR 1.88, p=0.059) (**Table 3**). When the multivariable analysis was adjusted to only include variables that were statistically significant (p<0.05) on the univariable analysis, age >65 years (p=0.0010), mitoses ≥1/mm^2^ (p=0.0492) and ulceration (p=0.0372) remained significant (**Supplement 2**).

**Table 3:**
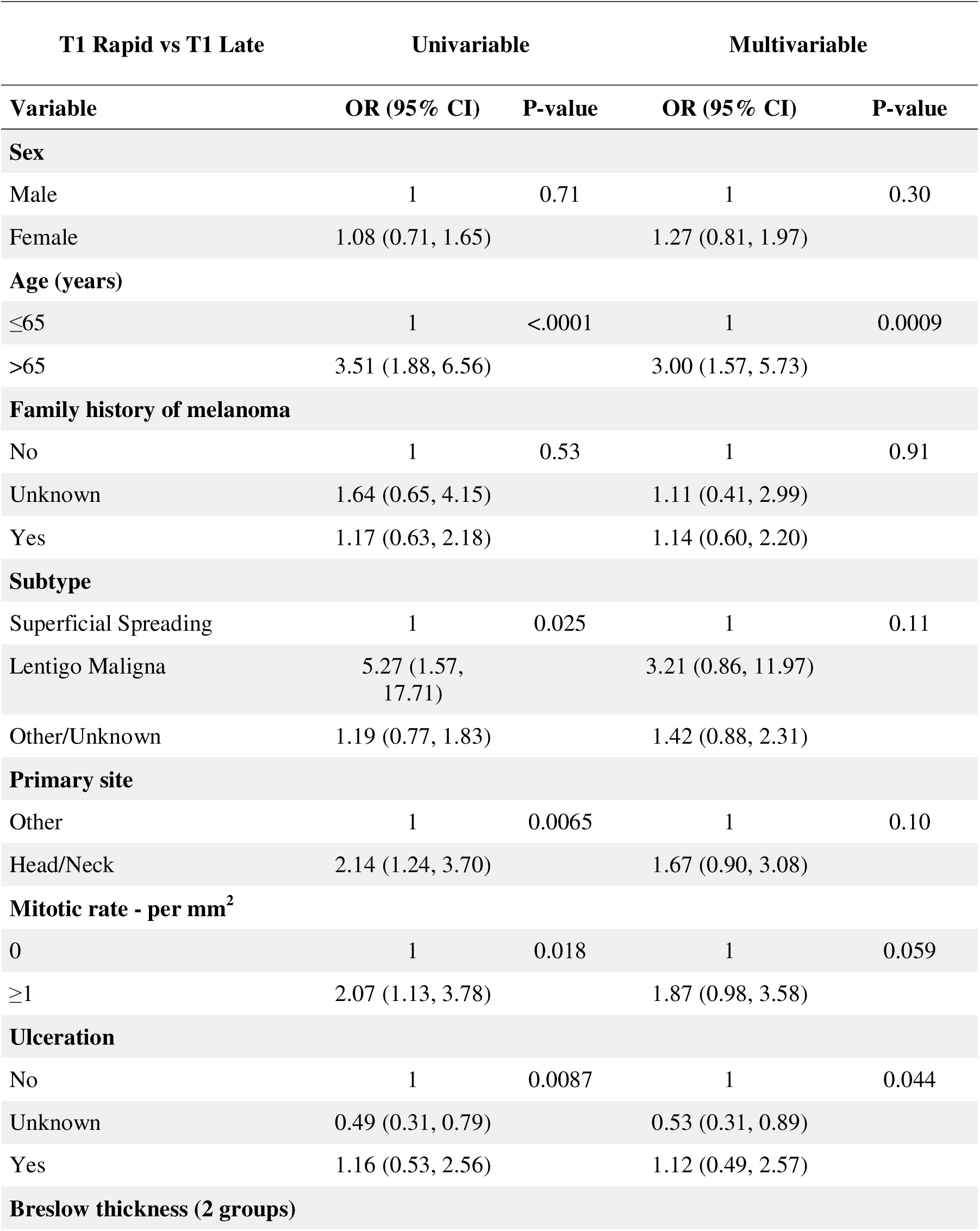
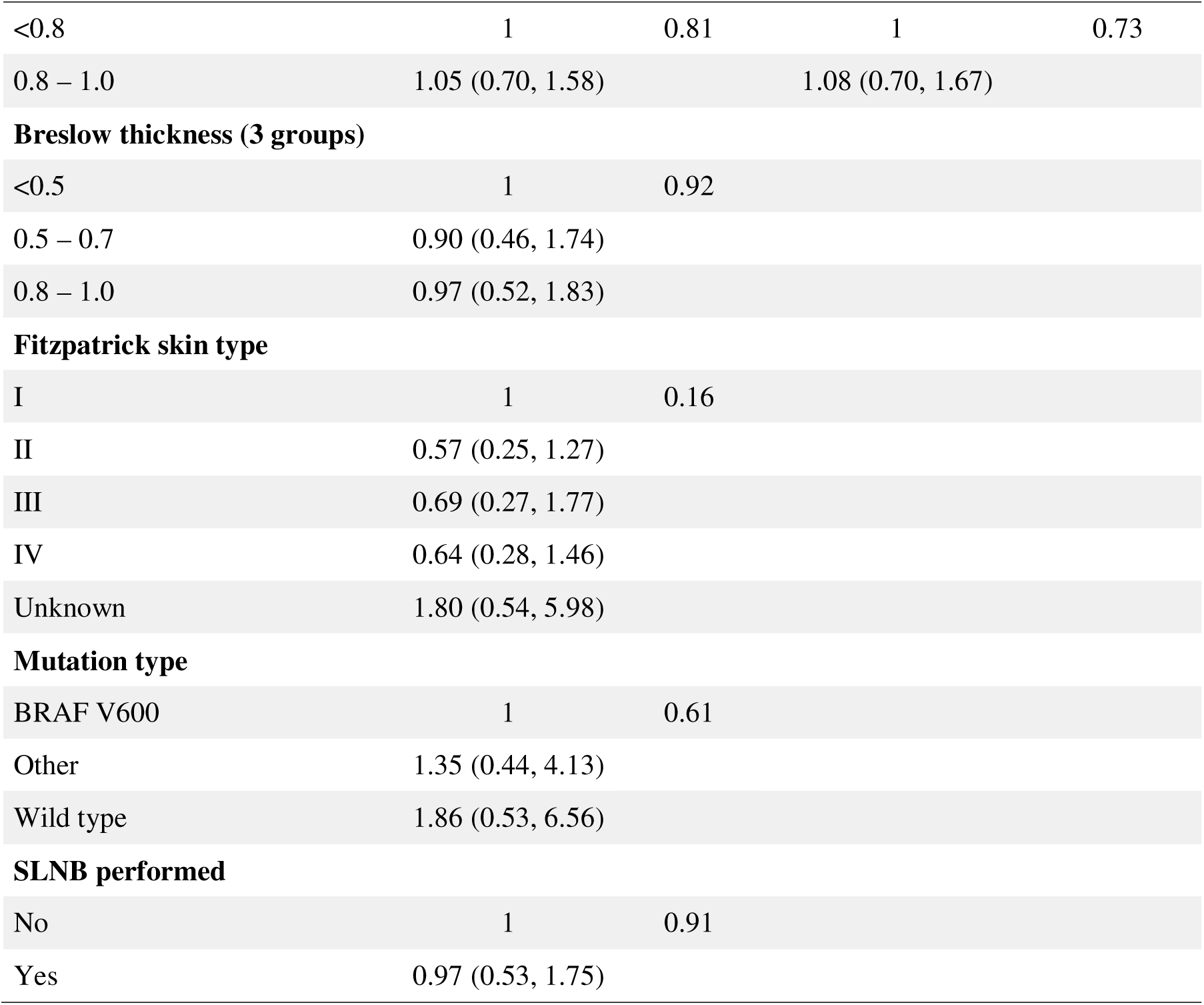
Univariable and Multivariable logistic regression analysis for factors associated with first recurrence, on pairwise testing comparing T1 early recurrences (event outcome, 1) with T1 late recurrences (non-event, 0). Variables that were statically significant (p<0.05) on univariable analysis or considered clinically significant were included in the multivariable analysis. SLNB, sentinel lymph node biopsy.

Age >65 yrs (p<0.0001), head or neck primary site (p=0.008) and mitoses ≥1/mm^2^ (p=0.043) were significantly associated with T1 rapid recurrence compared to T1 late recurrence (χ^2^ test). Furthermore, age >65 yrs combined with head or neck primary site (p=0.0028) and mitoses ≥1/mm^2^ combined with non-head or neck primary (p=0.0052) were significantly associated with T1 rapid recurrence.

## Discussion

To our knowledge, this is one of the largest studies to explore the characteristics of thin, T1 melanoma with rapid recurrence of disease within 2 years of diagnosis. Furthermore, this study is unique in that it also characterises thin, T1 melanoma with very late recurrence, occurring at least 10 years since the primary melanoma diagnosis, with a median follow up of over 17 years.

We found that several clinical and pathological features are associated with rapid recurrence compared to late recurrence of T1 melanomas, including age >65 years, LM histological subtype, head or neck primary site, increased mitotic rate and ulceration. Indeed, older age, increased mitotic rate and ulceration have previously been shown to be factors associated with poorer prognosis in patients with advanced melanoma.^23,25^ Head and neck primary site has also demonstrated a correlation with poorer prognosis compared to other sites, despite higher UV exposure at this location resulting in a subsequent higher tumour mutational burden (TMB).^26–28^ More recent studies have shown a particularly high responsiveness to PD-1 inhibition for head and neck melanomas. This suggests that it is the molecular biology of these tumours that determines prognosis and predicts responsiveness to selected therapies.^26–28^ Overall, this study therefore demonstrates that these predictors of poor prognosis in later stage melanoma are also implicated in rapid recurrence of early-stage melanoma.

Where data was available, the majority (59%) of patients in this study noted that their primary melanoma developed from a change in an old lesion rather than a new lesion **(Supplement 1)**. This was particularly noticeable in the late recurrence group where 82% of patients reported that their primary melanoma developed from an old skin lesion rather than a new lesion, noting however that data was not captured for many patients. Nevertheless, this finding may reflect unique underlying biology of melanomas that arise in longstanding skin lesions and then recur over a decade after removal of the primary, perhaps due to an inherently indolent nature or strong interaction with the host immune system.^29,30^ Furthermore, a significant difference was observed in the pattern of recurrence between the rapid and late groups, with the majority of recurrences in the rapid group being symptomatic and locoregional disease, compared to the majority of recurrences in the late group being asymptomatic and metastatic (p=0.0024 and p<0.001, respectively). This may reflect differences in underlying biology between the two groups, as we hypothesised. Additionally, despite these differences in recurrence patterns between the two T1 groups, the death rate from melanoma was similar between the two groups (53% T1 rapid vs 61% T1 late, p=0.23) likely reflecting aggressive biology in the T1 rapid group.

It is well established that increased BT of a primary melanoma is associated with worse prognosis.^23^ Because of this, there is a widespread perception that most melanoma deaths result from thick primaries. However, most patients diagnosed with melanoma each year have stage I melanoma.

Whilst the absolute risk of recurrence and death from stage I melanoma remains low, early-stage melanoma results in the major contribution to total mortality, due to the high number of cases. Indeed, long term follow up has shown that after 30 years, more melanoma related deaths occur due to T1 melanoma than T4 melanoma.^4^

As such, there is an urgent need to develop tools and strategies that can help clinicians identify patients with thin, T1 melanoma that are at high risk of recurrence. Whilst we have identified several clinical and pathological characteristics associated with rapid recurrence in this study, further translational research, such as using spatial transcriptomics and other novel multiomic technologies, is needed to identify predictive biomarkers for early-stage melanoma recurrence. By stratifying patients by level of recurrence risk, management can be tailored accordingly. For example, patients at high risk of recurrence may be more aggressively managed with wider margins of excision and sentinel lymph node biopsy (SLNB) and may undergo more rigorous surveillance imaging and follow up. Furthermore, as systemic therapy has moved earlier in the treatment paradigm for melanoma-from palliative treatment of stage IV disease to now neoadjuvant therapy in resectable disease-it is possible that, in the future, some patients with stage I disease may also benefit from adjuvant systemic therapy.

A significant amount of work is being undertaken in this field, with several GEP panels developed to identify patients with early-stage melanoma at high risk of recurrence. For example, the DecisionDx-Melanoma platform by Castle Biosciences uses a 31-gene GEP to predict an individual’s risk of metastasis or recurrence, as well as the chances of having a positive sentinel lymph node biopsy.^31,32^ Whilst a number of GEP panels have been commercialised and are available at a cost in some countries such as the US, it is unclear whether such platforms provide additional prognostic information beyond the AJCC staging system and established key clinico-pathological features.^33,34^ Indeed, international treatment guidelines clearly state that commercially available GEP tests are not recommended to be incorporated into routine melanoma care and that ‘use of such tests requires further prospective investigation in large, contemporary datasets of unselected patients’.^35^ Thus, the impact of GEP panels on treatment recommendations and overall clinical utility is not yet established. Furthermore, given that patients with thin melanoma experience 10-year survival rates of over 95%, a prognostic test or model would need to have excellent performance accuracy to be brought into routine clinical practice.

This study highlights the importance of long-term follow-up assessment of patients with T1 melanoma, as a large number of patients with recurrence after 10 years of diagnosis were able to be included in this study. This is despite significant limitations in record keeping and long-term follow-up of such patients noted by almost all participating sites. Indeed, where data on method of recurrence detection was available, the majority (74%) of patients in the T1 late recurrence group had their recurrence detected via clinical examination or imaging, demonstrating the importance of long term clinical follow up of these patients. It has been shown that late mortality due to melanoma is more common than early mortality in T1 melanoma, and this is reflected in our study where the majority of patients in the late recurrence group presented with stage IV disease, compared to the majority of patients in the early recurrence group having stage III disease.^4,20^ However, almost all surveillance programs in melanoma units will not capture these late and advanced stage recurrences, as they end after 10 years of follow-up duration. Furthermore, they are largely focussed on detection of a secondary primary melanoma rather than recurrence of a previous low-risk melanoma and have low long term compliance rates. Clinicians and innovative systems need to develop to be able to follow and monitor such low-risk patients in the long term, beyond 10 years, in an accurate and cost-effective manner.

Limitations of this study include the retrospective nature of the data collection and associated biases. The majority of patients did not have data captured on TILs, regression and LVI in the primary melanoma, reflecting the limitations of retrospective data collection especially in the context of early-stage melanoma. Indeed, over 20 additional sites globally were approached to participate in this study but were unable to do so due to the absence of a robust database capturing details on early-stage melanoma cases at their site. We also note that, particularly in the T1 late recurrence group, there is potential for the documented melanoma recurrence to be due to a second primary melanoma rather than the index lesion. Some inherent biases are evident due to the nature of the patient groups of interest in this study; for example, it is not surprising that the majority of patients in the T1 rapid group were male, as male sex is known to be associated with more aggressive melanoma. Furthermore, the finding that older age is associated with early rather than late recurrence may be explained by the lessened likelihood that older patients will survive long enough to reach the 10 year mark required to qualify as a late recurrence. The inclusion of a comparator cohort may have allowed for this issue to be addressed. Whilst there is potential for a bias in the OS analysis as patients in the late recurrence group may be more likely to receive new therapies, a significant difference in systemic therapies received between the two groups was not observed (**Table 2**). Lastly, whilst this study identifies factors associated with rapid and late recurrence in early-stage melanoma, it does not identify the percentage of all early-stage patients who have these risk factors, as patients were identified for inclusion in this study based on their future outcome already being known.

In conclusion, rapid recurrence of thin, T1 melanoma within 2 years of diagnosis is strongly associated with age >65 years, lentigo maligna melanoma histopathological subtype, head or neck primary melanoma site, mitoses ≥1/mm^2^ and ulceration. Patients harbouring these characteristics at melanoma diagnosis may benefit from more aggressive management and closer surveillance to enable timely detection of recurrence. Furthermore, patterns of disease recurrence vary between T1 melanoma rapid and late recurrence, suggesting differences in underlying biology. Further mechanistic studies are needed to explore these findings.

## ADDITIONAL INFORMATION

## Acknowledgements

Stacey Stern, Manager, Data Management, St John’s Cancer Institute, California, USA

· Christian U Blank, Department of Medical Oncology, Netherlands Cancer Institute; Department of Medical Oncology, Leiden University Medical Centre.

· Isabell Pieper Scholz and Marc Scherrer, Data Management, University Hospital Zurich

## Authors’ contributions

Study concept and design: PB, GAM. Data collection: PB. Data supply: all authors. Quality control of data and algorithms: PB. Data analysis and interpretation: PB, TW, SNL, ATP, GAM. Statistical analysis: PB, TW, SNL. Manuscript preparation: PB, ATP, GAM. Manuscript editing: PB, ATP, GAM. All authors revised the manuscript and approved the submission. All authors are agreeable to be accountable for all aspects of the submitted work.

## Ethics approval

Each individual site gained ethics approval with their local institutional ethics board.

## Patient consent to participate

Not required.

## Consent for publication

All authors consent to publication of this manuscript.

## Data availability

Data are available upon reasonable request. All data relevant to the study are included in the article or uploaded as supplemental information. Deidentified participant data are available from PB upon request.

