## Supplements for "Clinical and pathological characteristics of thin cutaneous melanomas with rapid recurrence"

**Supplement 1:** Baseline characteristics for all patients and stratified by recurrence groups, where characteristic had more that 25% of patients with unknown data. TILs, Tumour Infiltrating Lymphocytes; LVI, lymphovascular invasion; WLE, wide local excision.

| Patient and disease characteristics | Overall  (N = 442) | T1 recurrence  ≤2 yr  (N = 310) | T1 recurrence ≥10 yr  (N = 132) | P-value |
| --- | --- | --- | --- | --- |
| TILs |  |  |  |  |
| Absent | 43 (50.6%) | 38 (50.7%) | 5 (50.0%) | 0.99 |
| Present, non-brisk | 31 (36.5%) | 27 (36.0%) | 4 (40.0%) |  |
| Present, brisk | 8 (9.4%) | 7 (9.3%) | 1 (10.0%) |  |
| Present, not specified | 3 (3.5%) | 3 (4.0%) | 0 |  |
| Unknown | 357 | 235 | 122 |  |
| History of chronic sun damage |  |  |  |  |
| No | 24 (28.2%) | 18 (26.9%) | 6 (33.3%) | 0.59 |
| Yes | 61 (71.8%) | 49 (73.1%) | 12 (66.7%) |  |
| Unknown | 357 | 243 | 114 |  |
| Primary melanoma origin |  |  |  |  |
| Change in old lesion | 71 (59.2%) | 49 (52.7%) | 22 (81.5%) | 0.0074 |
| New lesion | 49 (40.8%) | 44 (47.3%) | 5 (18.5%) |  |
| Unknown | 322 | 217 | 105 |  |
| Primary melanoma diagnosed by |  |  |  |  |
| Patient noticed | 74 (63.2%) | 63 (67.7%) | 11 (45.8%) | 0.047 |
| Doctor noticed | 43 (36.8%) | 30 (32.3%) | 13 (54.2%) |  |
| Unknown | 325 | 217 | 108 |  |
| Regression |  |  |  |  |
| Absent | 142 (81.1%) | 115 (82.1%) | 27 (77.1%) | 0.83 |
| Early/TILs present | 5 (2.9%) | 4 (2.9%) | 1 (2.9%) |  |
| Intermediate/partial/focal | 14 (8.0%) | 10 (7.1%) | 4 (11.4%) |  |
| Late/complete | 2 (1.1%) | 2 (1.4%) | 0 |  |
| Present, not specified | 12 (6.9%) | 9 (6.4%) | 3 (8.6%) |  |
| Unknown | 267 | 170 | 97 |  |
| LVI |  |  |  |  |
| No | 120 (88.9%) | 94 (87.0%) | 26 (96.3%) | 0.30 |
| Yes | 15 (11.1%) | 14 (13.0%) | 1 (3.7%) |  |
| Unknown | 307 | 202 | 105 |  |
| Associated naevus |  |  |  |  |
| No | 69 (60.0%) | 60 (64.5%) | 9 (40.9%) | 0.042 |
| Yes | 46 (40.0%) | 33 (35.5%) | 13 (59.1%) |  |
| Unknown | 327 | 217 | 110 |  |
| Extent of WLE |  |  |  |  |
| 1cm | 97 (74.6%) | 72 (69.2%) | 25 (96.2%) | 0.016 |
| 2cm | 18 (13.8%) | 18 (17.3%) | 0 |  |
| Other | 9 (6.9%) | 9 (8.7%) | 0 |  |
| Not done | 6 (4.6%) | 5 (4.8%) | 1 (3.8%) |  |
| Unknown | 312 | 206 | 106 |  |

**Supplement 2:** Univariable and Multivariable logistic regression analysis for factors associated with 1^st^ recurrence, on pairwise testing comparing T1 early recurrences (event outcome, 1) with T1 late recurrences (non-event, 0). Only variables that were statically significant (p<0.05) on univariable analysis were included in the multivariable analysis. SLNB, sentinel lymph node biopsy.

| **T1 Rapid vs T1 Late** | **Univariable** | | **Multivariable (Ulceration only)** | |
| --- | --- | --- | --- | --- |
| **Variable** | **OR (95% CI)** | **P-value** | **OR (95% CI)** | **P-value** |
| **Sex** |  |  |  |  |
| Male | 1 | 0.71 |  |  |
| Female | 1.08 (0.71, 1.65) |  |  |  |
| **Age (categorised)** |  |  |  |  |
| <=65 | 1 | <.0001 | 1 | 0.0010 |
| >65 | 3.51 (1.88, 6.56) |  | 2.94 (1.55, 5.58) |  |
| **Family history of melanoma** |  |  |  |  |
| No | 1 | 0.53 |  |  |
| Unknown | 1.64 (0.65, 4.15) |  |  |  |
| Yes | 1.17 (0.63, 2.18) |  |  |  |
| **Subtype** |  |  |  |  |
| SSM | 1 | 0.025 | 1 | 0.11 |
| LM | 5.27 (1.57, 17.71) |  | 3.16 (0.85, 11.76) |  |
| Other/Unknown | 1.19 (0.77, 1.83) |  | 1.42 (0.88, 2.29) |  |
| **Primary site** |  |  |  |  |
| Other | 1 | 0.0065 | 1 | 0.13 |
| Head/Neck | 2.14 (1.24, 3.70) |  | 1.59 (0.87, 2.93) |  |
| **Mitoses** |  |  |  |  |
| <1 | 1 | 0.018 | 1 | 0.049 |
| ≥1 | 2.07 (1.13, 3.78) |  | 1.91 (1.00, 3.63) |  |
| **Ulceration** |  |  |  |  |
| No | 1 | 0.0087 | 1 | 0.037 |
| Unknown | 0.49 (0.31, 0.79) |  | 0.52 (0.31, 0.88) |  |
| Yes | 1.16 (0.53, 2.56) |  | 1.12 (0.49, 2.55) |  |
| **Breslow thickness (2 groups)** |  |  |  |  |
| <0.8 | 1 | 0.81 |  |  |
| 0.8 – 1.0 | 1.05 (0.70, 1.58) |  |  |  |
| **Breslow thickness (3 groups)** |  |  |  |  |
| <0.5 | 1 | 0.92 |  |  |
| 0.5 – 0.7 | 0.90 (0.46, 1.74) |  |  |  |
| 0.8 – 1.0 | 0.97 (0.52, 1.83) |  |  |  |
| **Fitzpatrick skin type** |  |  |  |  |
| I | 1 | 0.16 |  |  |
| II | 0.57 (0.25, 1.27) |  |  |  |
| III | 0.69 (0.27, 1.77) |  |  |  |
| IV | 0.64 (0.28, 1.46) |  |  |  |
| Unknown | 1.80 (0.54, 5.98) |  |  |  |
| **Mutation type** |  |  |  |  |
| BRAF v600 | 1 | 0.61 |  |  |
| Other | 1.35 (0.44, 4.13) |  |  |  |
| WT | 1.86 (0.53, 6.56) |  |  |  |
| **SLNB performed** |  |  |  |  |
| No | 1 | 0.91 |  |  |
| Yes | 0.97 (0.53, 1.75) |  |  |  |
